# SputNet: expert-level deep learning differential cell counts across whole respiratory cytospins expose the limits of manual counting

**DOI:** 10.64898/2026.11.26.26369019

**Authors:** Shayan Rasaratnam, Gökhan Yildirim, Yuliya V Karpievitch, Jodie L Simpson, Luke W Garratt

**Author notes:** Corresponding to: Shayan Rasaratnam,; Luke Garratt.

## Abstract

**Background:** Although differential cell counts provide clinically actionable biomarkers of airway inflammation, labour intensity limits routine clinical use. We developed SputNet, a deep learning approach to the problem. We tested whether SputNet could count with agreement comparable to that between expert cytologists. Using whole cytospin analysis by SputNet, we also investigated how often human subset counts may disagree with the whole-slide result.

**Methods:** We trained a dual-step deep learning pipeline (object detection then classification) on 59,974 cells from 3,678 development images of asthma, COPD or cystic fibrosis cytospins. Model performance was evaluated on a locked test set of 600 images independently annotated by four cytologists, using an agreement score based on the F1 metric. SputNet was applied to 191 whole cytospins to map spatial heterogeneity and simulate conventional human subset counts.

**Results:** Inter-cytologist agreement was 0.71 for immune cells, but poor for eosinophils (0.51) and lymphocytes (0.26). SputNet’s overall agreement was 0.74 (cytologists 0.70–0.72) and met or exceeded the human mean for every class. SputNet counted a mean of 22,372 cells per whole cytospin in a mean time of 16 minutes. When whole-slide results were near eosinophilia or neutrophilia cut-offs, simulated human subset counts often gave a different determination, even for counts of up to 1000 cells.

**Conclusions:** SputNet delivers human-level sputum cell classification and whole-slide cytology that avoids the sampling variability of human subset counts. Multicentre external validation is needed before integration into respiratory clinical care.

## Introduction

Artificial Intelligence (AI), which includes machine learning (ML) and computer vision (CV), is demonstrating remarkable abilities to transform clinical decision-making across diverse medical fields (1). Respiratory differential cell counts measure airway cellular inflammation, but automated counting of these preparations through AI remains underexplored. Neutrophil and eosinophil counts in particular serve as biomarkers for inflammatory subtype, disease severity, and treatment responsiveness, across a broad range of chronic respiratory diseases (2). These neutrophil and eosinophil counts inform treatment strategies and improve clinical outcomes in asthma (3, 4) and can lower asthma-related healthcare expenses by 19 percent (5). While this economic benefit is best characterised in asthma, emerging evidence supports similar clinical utility in chronic obstructive pulmonary disease (6) and cystic fibrosis (7).

Despite clinical utility advantages, widespread clinical utilisation of respiratory cytospins remained limited due to manual differential cell counting being time-consuming, labour-intensive, and subject to inter-observer variability (8). The process requires typically involving manual counting of 300 to 600 total cells per slide (9, 10) as although 10,000-40,000 cells are usually loaded onto a cytospin, counting all cells is impractical. A trained cytologist therefore counts a subset, typically in a lawn-mower pattern, until a target total is reached. Human error, which can occur due to inconsistent cell classification, errors of omission, or selection bias regarding the area to be analysed, also introduces undesired variability (9). These errors can skew immune cell proportions and will particularly affect eosinophilic inflammation diagnosis, as these cells are a minor proportion in healthy airways. For example, current guidelines defining eosinophilic inflammation beyond normal ranges as ≥2.3% of eosinophils (11), correspond to ≥9.2 eosinophils in a standard 400 cell count. Observer error and/or uneven distribution of eosinophils on the cytospins could influence the diagnosis of eosinophilic inflammation and potential treatment decisions. Remedial solutions of recounts or secondary independent assessments only impose further time and cost barriers.

These challenges could all be addressed by automated cell identification and counting. Existing automated systems such as CellaVision DM96 (12) rely on specific imaging platforms, constraining their broad clinical applicability. To address this, we previously introduced DCNet, a model trained solely on centre-point-annotated images generated by agreement of multiple annotators, which achieved human-level performance (13). In this study, we present SputNet, which builds on DCNet through a larger annotated dataset of respiratory cytospins and a dual-step architecture that improves detection of individual cells on dense slides, while incorporating cell size, shape, and colour features. With SputNet able to analyse entire cytospins preparations, we also explored whole-slide data to demonstrate spatial heterogeneity of cell distribution and its effect on variability in human-based approaches that count subsets of the slide area.

## Methods

### Study design and reporting

This retrospective study describes the development and held-out evaluation of a deep learning model for detection and classification of cells in archived respiratory cytospins. Reporting of model development and evaluation was guided by the applicable items of TRIPOD+AI (14). A completed TRIPOD+AI checklist is provided in the supplement. A separate study protocol publication was not prepared, nor was the study prospectively registered.

### Cytospin data and annotations

Respiratory sample cytospins were sampled from archives of four research studies of airway inflammation in respiratory disease, contributed by three tertiary research institutions. Cytospins were collated and imaged in this study under University of Western Australia HREC approval 2023/ET000729. Additional consent was not sought as these archived samples were all consented at the time of collection for the quantification of immune cells including differential count. Overall, all cytospins underwent routine protocols for cytospin generation of respiratory cells (10), further details are provided in the supplementary material. To ensure a robust training set, cytospins were retrieved from study archives with a selection emphasis to represent the range of observed cell densities, percentage of neutrophils, percentage of eosinophils, and percentage of lymphocytes, as well as samples that were considered challenging to count by the original study investigators. All cytospins were digitally imaged under 100x oil magnification by the Centre for Microscopy, Characterisation and Analysis (University of Western Australia) using a ScanScope OS (Leica Biosystems, Australia).

Areas of images were selected randomly in ImageScope (Leica Biosystems, Australia), exported and uploaded to LabelBox for annotation. Cells were manually annotated by trained cytologists as eosinophils, lymphocytes, macrophages/monocytes, neutrophils, epithelial cells, or unknown cells, marking the centre-point of the cell. Bounding boxes were generated by taking the average diameter of a specific cell class, based on our previously reported data (13).

A total of 4,278 images from 101 cytospins were annotated, by four cytologists, each with >15 years of experience in respiratory differential cell counts. The development set comprised 3,678 images from 71 cytospins and was used for all model training and selection through four-fold, cytospin-grouped cross-validation, in which every image from a given cytospin was kept within the same fold. Consensus annotations for the development set were created from four cytologists. A separate test set of 600 images from 30 cytospins was locked before model development and was not accessed during training, hyperparameter selection or threshold calibration. The test set was independently annotated by all four cytologists with no prior knowledge of the other cytologists’ annotations; cytologists could also label an object as an unknown cell where a confident classification was not possible. These four independent annotation sets were used to assess both inter-annotator agreement and SputNet accuracy.

The development set consisted of 59,974 consensus-classified immune cells (3,817 eosinophils, 4,338 lymphocytes, 27,528 macrophages, 24,291 neutrophils), plus 3,588 columnar and 69 squamous epithelial cells, and 5,835 unknown cell objects used only as cell detection targets. Cell counts for each cytologist on the 600 test images are provided in the supplementary material.

### Model architecture

SputNet employs a two-step machine learning architecture (Figure 1), replacing the heatmap-based semantic segmentation approach used in DCNet. First, cell detection is performed using a Faster R-CNN object detector (15) to predict a bounding box and confidence score for each individual cell (16). Second, the model employs EfficientNetV2, an image classifier (17), for cell identification as either eosinophils, lymphocytes, macrophages, neutrophils, or columnar or squamous epithelial cells. Both models were trained independently and further details on these models are provided in the supplementary material.

**Figure 1.**
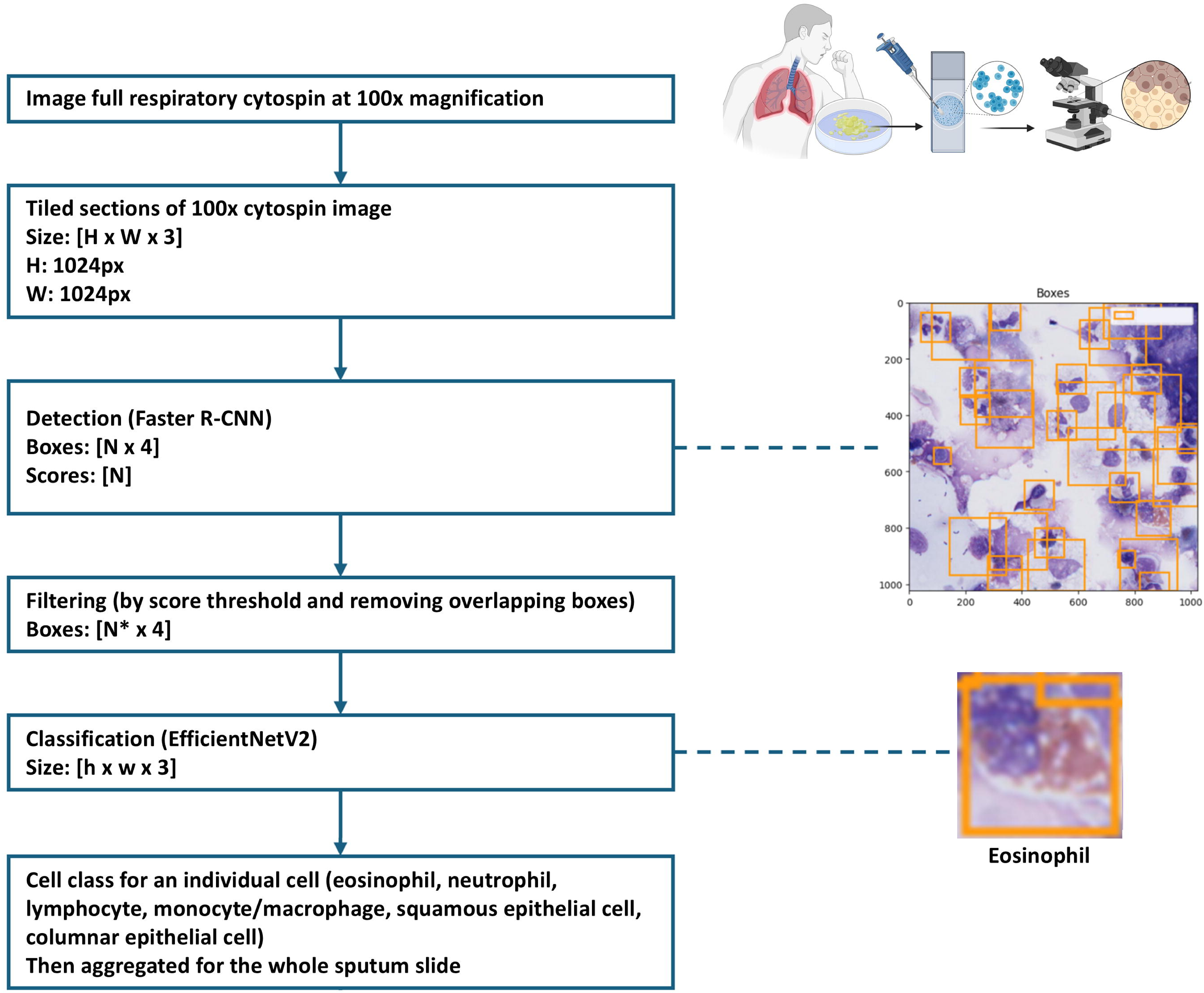
Block diagram of SputNet pipeline. Cytospins generated from respiratory samples were digitised through 100x scan imaging. Scans were split into 1024 pixel x 1024 pixel tiles, leading to ~4,875 tiles per slide. Each individual tile then followed a two-step pipeline. Object detection (Faster R-CNN) was used to detect cells, followed by filtering cells by probability score and overlap, and then cell-type classification (EfficientNetV2) producing label across the major cell types observed in respiratory samples.

### Measuring performance

Manual sputum cell counts by trained cytologists are the current gold standard. We evaluated SputNet against human annotators, acknowledging that discrepancies between human assessors do occur, especially with rarer cell types like eosinophils and lymphocytes, and no ground truth may exist for some cell objects.

We first measured agreement at the cell level between human annotators, then we compared SputNet’s agreement at the cell level with annotators. This was performed on the 600 test images (30 cytospins), which contained between 6,183 and 9,241 immune cells based on the annotator. Agreement between two annotators (for example A and B) was measured using the “agreement score”, defined as the mean of the F1 score of A with respect to B and the F1 score of B with respect to A.

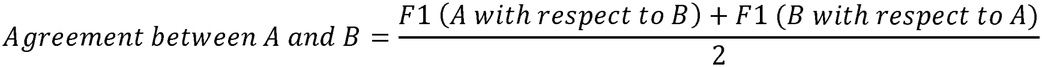

The F1 score was chosen over precision or recall because it penalises high rates of both false positives *and* false negatives. For this reason, F1 score is commonly used to assess performance of image classification models (18). To determine the F1 score of A with respect to B, we assume B’s annotations are true, and then A’s annotations are compared with B to assess the rate of true positives, false positives, and false negatives. These values can then be inputted into the following equation:

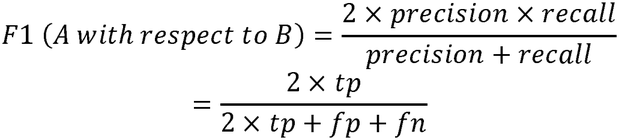

An F1 agreement score close to 1 indicates excellent agreement, while a score near 0 indicates total disagreement.

### Heterogeneity analyses

Whole-slide SputNet output (tile coordinates and per-class counts) was analysed in R using RStudio™, details are provided in the supplementary material. Compositional diversity was summarised by Berger–Parker dominance, Gini– Simpson index, Pielou evenness, Shannon entropy and Hill numbers of order 1 and 2. Spatial unevenness of each class was summarised by the coefficient of variation (CoV) of tile-level proportions and spatial autocorrelation by global Moran’s I. To recreate the standard manual subset count performed by humans, lawn-mower trajectories were simulated across each whole-slide map until a target total (300, 400, 600 or 1000 cells) was reached, and each subset eosinophilia or neutrophilia call was compared with the whole-slide call

### Patient and public involvement

Patients and the public were not involved in the design, conduct, reporting or dissemination plans of this research.

## Results

A total of 191 cytospins from 140 individuals were used in this study. Respiratory presentations included asthma (n=32 induced sputum cytospins from 15 individuals), COPD (n=57 induced sputum cytospins from 55 individuals), CF (n=84 bronchoscopy cytospins from 52 individuals) and investigative bronchoscopies (n=18 bronchoscopy cytospins from 18 individuals). Of these, 101 cytospins were annotated: 71 cytospins formed the development set and 30 cytospins formed the locked test set. The remaining 90 cytospins were used only in the whole-slide analysis.

The mean overall agreement score between four human annotators was 0.711 ±0.019 for immune cells (Table 1). Agreement was poorest for lymphocytes (0.264 ±0.106) and was also moderate for eosinophils (0.510 ±0.056), which shows how observer variation could affect the diagnosis of eosinophilic inflammation. Cytologists frequently assigned different classes to the same object (Figure 2). SputNet’s agreement scores were higher than the cytologist mean for every class (eosinophils 0.52, lymphocytes 0.30, macrophages/monocytes 0.72, neutrophils 0.81, epithelial cells 0.37), and its overall agreement (0.74 for immune cells; 0.72 for all known cells) was numerically higher than that of each individual cytologist (0.70–0.72 and 0.68– 0.70, respectively).

**Table 1.**
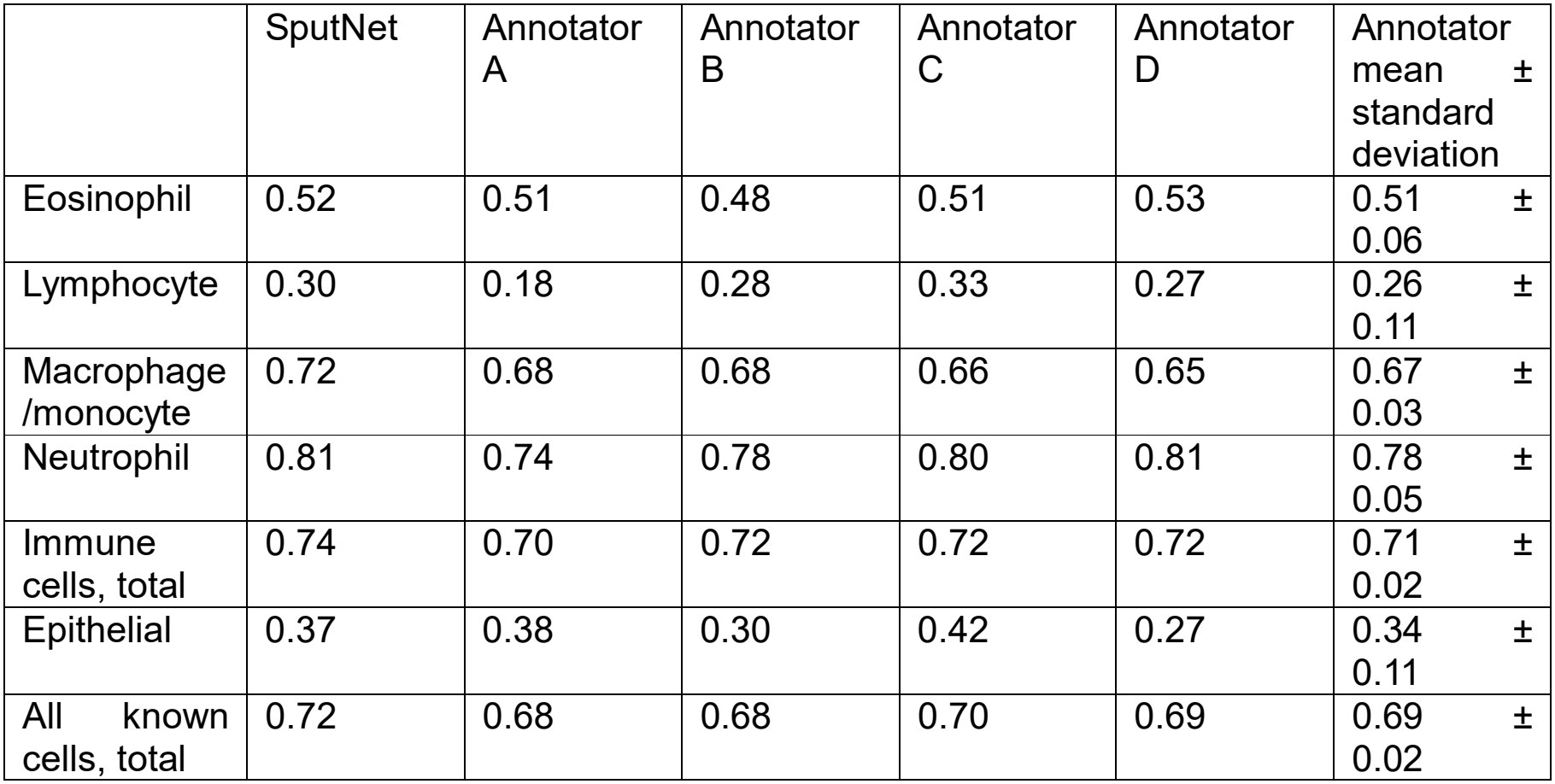
Mean agreement scores of SputNet and cytologists. Data from the locked test set only (600 images from n=30 cytospins). Agreement score = mean of the two directional F1 scores between a pair of annotators. SputNet’s scores were calculated against all four cytologists; each cytologist’s against the other three. Epithelial combines squamous and columnar epithelial cells. Immune cells: eosinophils, lymphocytes, macrophages/monocytes and neutrophils; all known cells: immune plus epithelial cells, excluding objects labelled unknown. SputNet’s scores were determined with respect to the human annotators, while the scores of the human annotators A-D were determined with respect to the other human annotators only.

|  | SputNet | Annotator A | Annotator B | Annotator C | Annotator D | Annotator mean ± standard deviation |
|---|---|---|---|---|---|---|
| Eosinophil | 0.52 | 0.51 | 0.48 | 0.51 | 0.53 | 0.51 ± 0.06 |
| Lymphocyte | 0.30 | 0.18 | 0.28 | 0.33 | 0.27 | 0.26 ± 0.11 |
| Macrophage /monocyte | 0.72 | 0.68 | 0.68 | 0.66 | 0.65 | 0.67 ± 0.03 |
| Neutrophil | 0.81 | 0.74 | 0.78 | 0.80 | 0.81 | 0.78 ± 0.05 |
| Immune cells, total | 0.74 | 0.70 | 0.72 | 0.72 | 0.72 | 0.71 ± 0.02 |
| Epithelial | 0.37 | 0.38 | 0.30 | 0.42 | 0.27 | 0.34 ± 0.11 |
| All known cells, total | 0.72 | 0.68 | 0.68 | 0.70 | 0.69 | 0.69 ± 0.02 |

**Figure 2.**
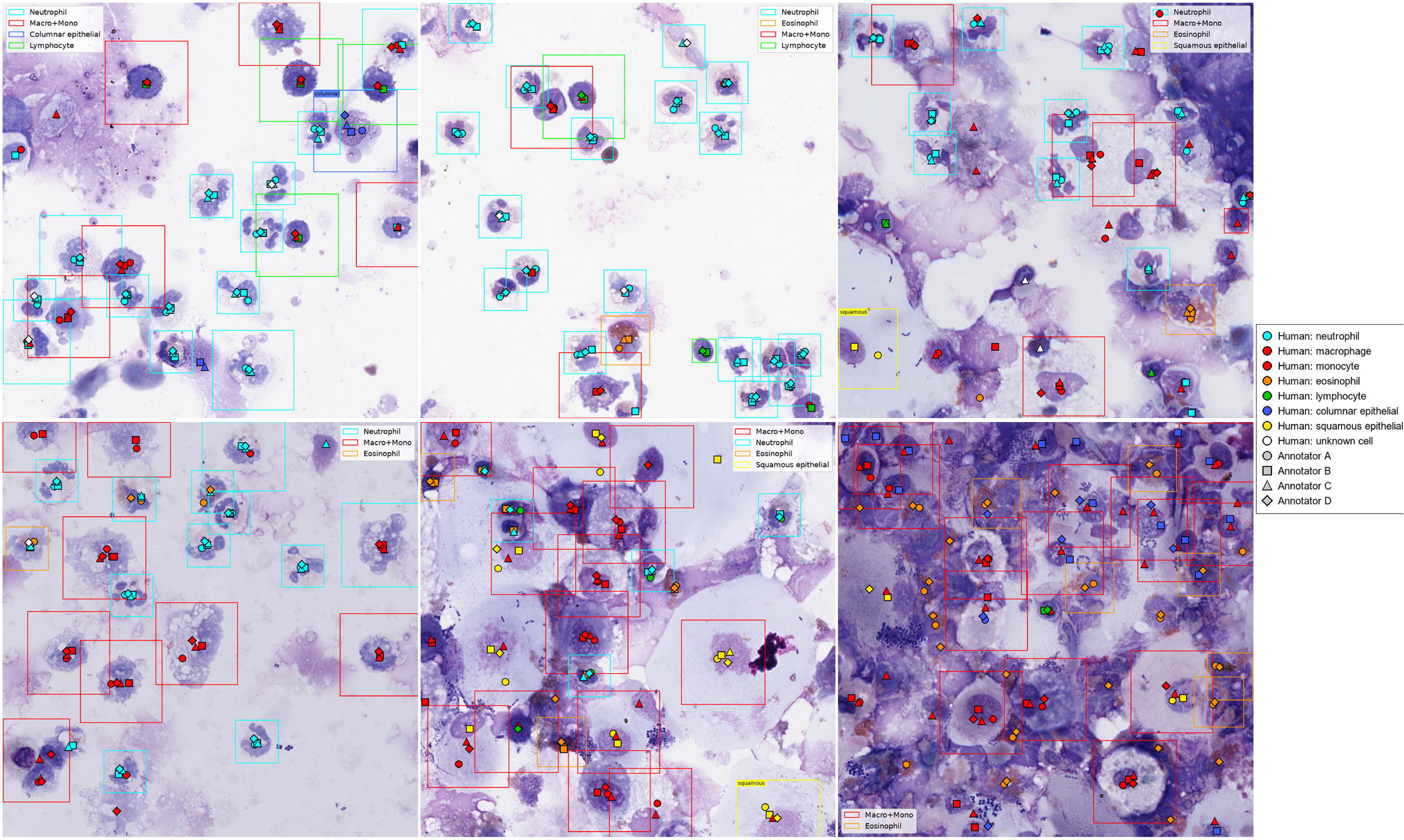
Example test-set tiles are from the locked test set showing independent annotations by four cytologists and SputNet predictions. Symbols mark cell centre points placed by each cytologist (symbol shape identifies annotators A–D; symbol fill identifies the assigned class). Boxes show SputNet detections, labelled by predicted class. Examples range from sparse, clean preparations (top) to dense, debris-rich preparations (bottom right).

SputNet typically assessed a single 100x image in 0.2 seconds and required a mean of 16 minutes to count a whole cytospin (median 4,875 images per cytospin; n=191 cytospins). Human annotators, by contrast, count 400-1000 cells. We used these whole-slide outputs (Figure 3A) to map cells across each slide (Figure 3B-E). Mean cell content was 22,372 per cytospin (SD 27,036; Figure 4A). Global heterogeneity metrics showed wide variation in dominance and evenness (Figure 4B). Eosinophils were particularly unevenly distributed (Figure 4C, D) but showed little local spatial autocorrelation, whereas neutrophils often formed clusters (Figure 4E). These patterns show why a human subset count can be influenced by where on the slide the count is being performed.

**Figure 3.**
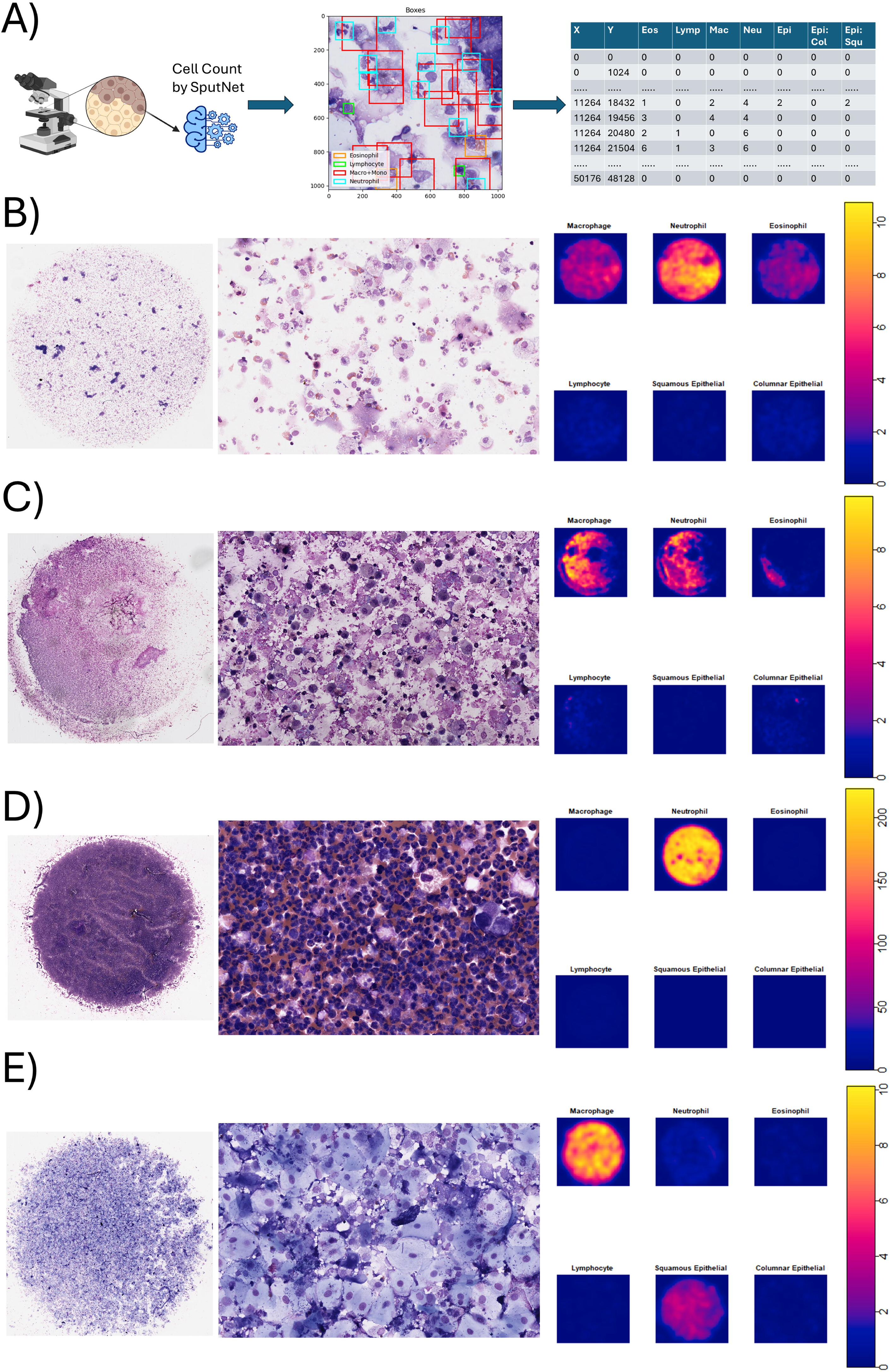
Whole-slide spatial mapping of cytospins by SputNet. (A) The x–y position and per-class cell counts of each 1024x1024 tile were generated by SputNet. (B–E) Whole-slide maps were built using the spatstat package in R, with four example cytospins shown across whole-slide image (left), representative 100× field (centre) and smoothed density maps for each cell class (right; scale represents cells per tile). Examples chosen to highlight (B) a mixed macrophage/neutrophil/eosinophil sample, (C) preparation with uneven clustering of classes, (D) a dense neutrophilic sample and (E) a macrophage-dominant preparation with substantial squamous epithelial cells.

**Figure 4.**
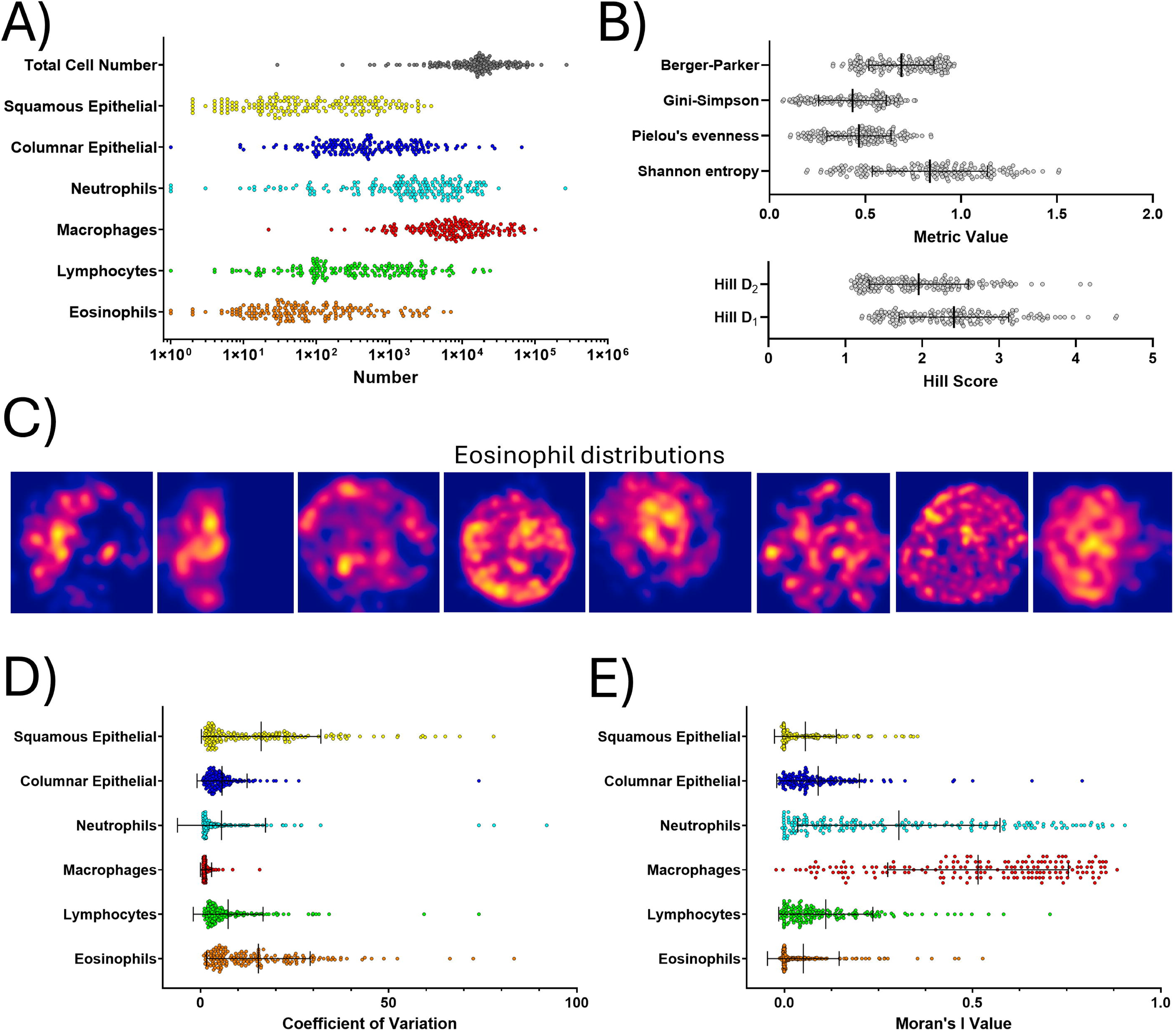
Cytospins from respiratory samples can be highly heterogenous in spatial composition. A) SputNet data summarised as number of cells on each cytospin, showing our set of cytospins featured a diverse range of cell densities and cell composition. B) The diversity of our cytospin set was further confirmed through multiple metrics of global balance between cell types. Berger-Parker measures the proportion of the single most abundant class, Gini-Simpson measures the probability that two randomly chosen objects belong to different classes, Pielou’s evenness measures how unbalanced the classes are, independent of total counts, and Shannon’s entropy measures the overall heterogeneity. Hill D_1_ measures the number of equally abundant classes and Hill D_2_ the number of dominant classes. C) Focusing on eosinophils, spatial distribution was often heterogenous. When we delve into this at a class level, considering proportions of cells, we can see that D) eosinophils do have the highest coefficient of variation in spatial distribution amongst immune cells. E) Moran’s I value, which measures autocorrelation of each class, shows that eosinophil distribution is random and not structured like neutrophils and macrophages. All data points represent an individual cytospin, with mean and SD shown.

We then compared simulated subset counts (Figure 5A) with the whole-slide classification. Whilst cytospins could vary greatly in the number of possible 400 cell counts (Figure 5B), which is influenced by the number of cells loaded during cytospin generation, we observed in our dataset that the median possible 400 cell counts on our cytospins was 114 (IQR 54-177). For each cytospin, we quantified the frequency with which manual 400 cell counts agreed with the whole-slide classification above or below the diagnostic cut-off (2.3% for eosinophils, 64.4% for neutrophils; Figure 5C-D). When the whole-slide result was between 1% to 6.5% eosinophils (Figure 5C), at least 1 in 8 simulated manual counts disagreed (e.g. false positive or false negative) for most cytospins, and for some cytospins disagreement was as high as 69% of manual counts. For neutrophilia, manual count classification was robust when the ground truth was below 55% or above 78% neutrophils (Figure 5D). Potential for incorrect results persisted when we simulated across small (300 cells, Figure 6A) and large target counts (600 cells, Figure 6B; 1000 cells, Figure 6C).

**Figure 5.**
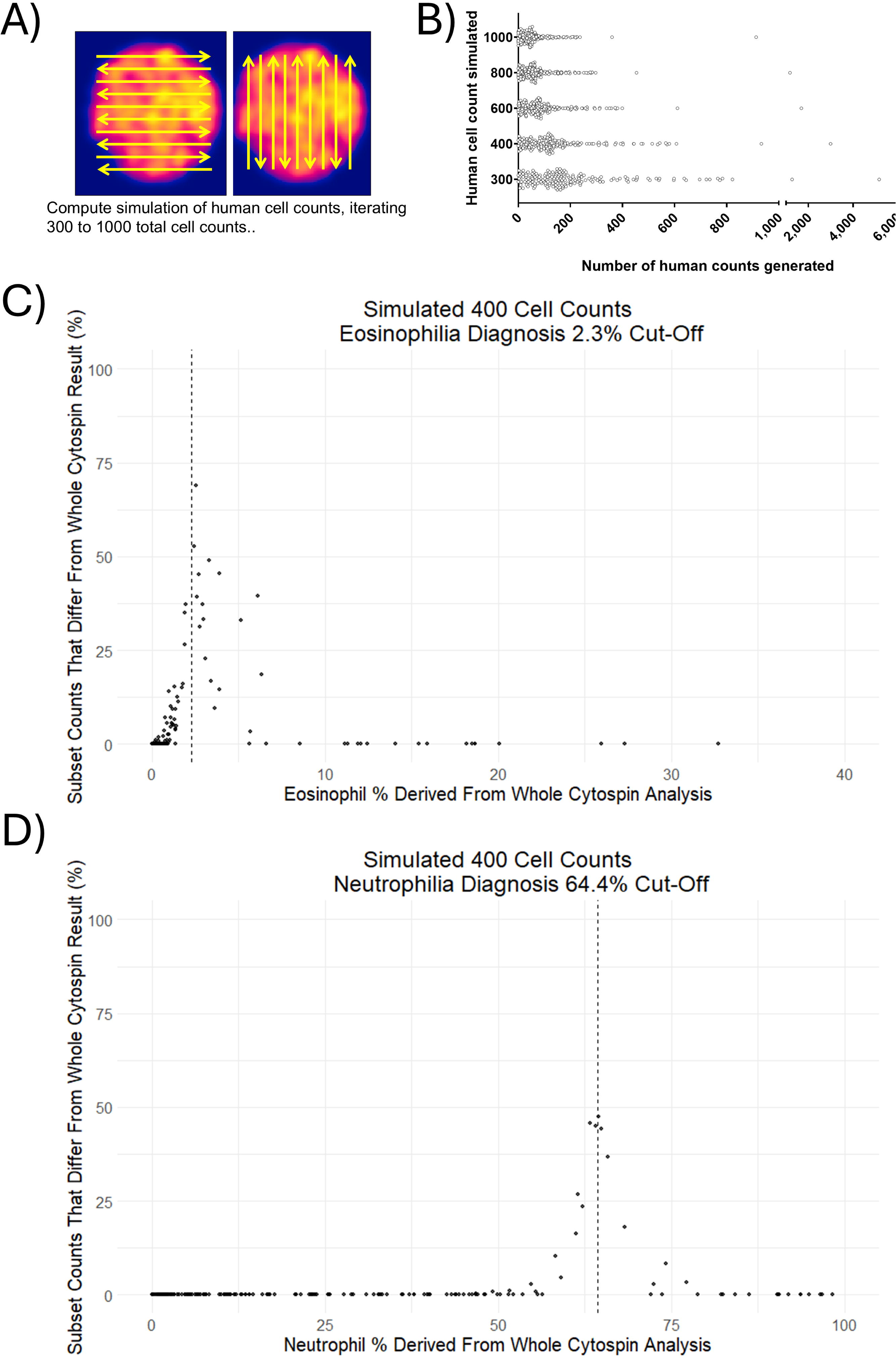
Heterogeneous distribution of cells across the cytospin means 400-cell subset counts can frequently disagree with the whole-slide classification. A) To simulate the typical human approach that counts a subset of the cytospins until target total cell number is reached, “lawn mower patterns” were simulated across the spatial data, across all directions. B) Number of distinct simulated counts per cytospin for targets of 300, 400, 600, 800 and 1000 cells. Each data point represents an individual cytospin, with mean number of subset counts and SD shown. (C, D) For each cytospin, the percentage of simulated 400-cell counts whose classification disagreed with the whole-cytospin classification, for (C) eosinophilia (≥2.3% eosinophils) and (D) neutrophils above the upper limit of normal (64.4%), plotted against the whole-cytospin percentage. Dashed lines mark the cut-offs. Each point is one cytospin.

**Figure 6.**
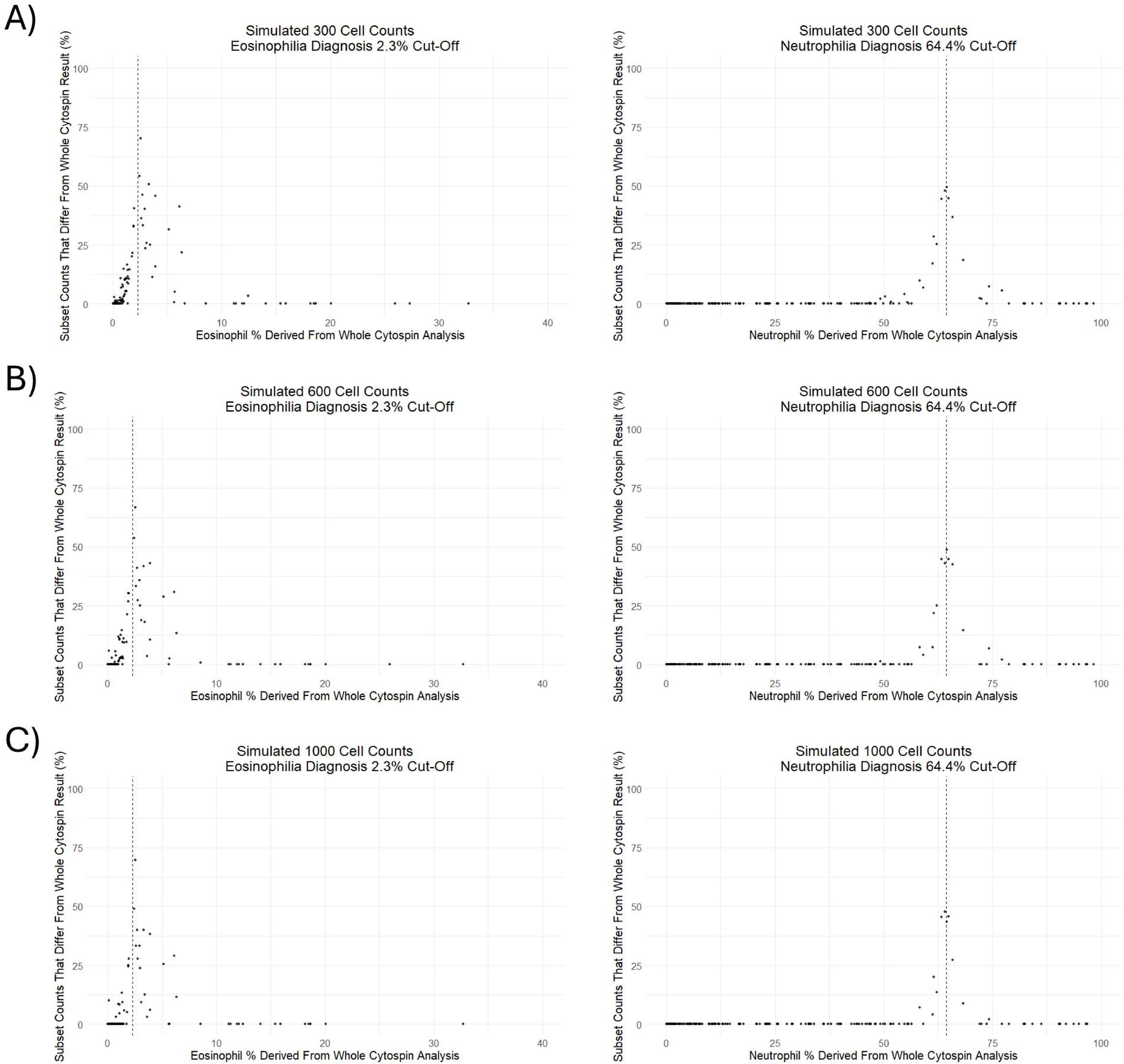
Disagreement persists with larger subset counts. Percentage of simulated counts disagreeing with the whole-slide classification for eosinophilia (left; 2.3% cut-off) and neutrophilia (right; 64.4% cut-off) at targets of (A) 300, (B) 600 and (C) 1000 cells, plotted against the whole-slide percentage. Dashed lines mark the cut-offs. Each point is one cytospin.

## Discussion

Our work demonstrates that AI-based respiratory cell count tools, exemplified by SputNet, can achieve performance comparable to four human experts. Unlike our previous study that primarily relied on precision alone (13), we assessed SputNet performance using an agreement score based on the F1 metric, a robust measure incorporating both false positives and negatives. Critically, SputNet had comparable agreement to experts for macrophages and neutrophils, the two most predominant cell types that most significantly affect differential count result calculations. Because our four cytologists assessed identical digital images, our inter-person data is more robust than an early study that measured agreement between cytologists after they counted the same slide through a microscope (9). Both SputNet and cytologists performed moderately for eosinophils and poorly with lymphocyte identification. Another automated approach, the CellaVision DM96, classifies preselected cells and also had modest sensitivity for eosinophilia and low sensitivity for neutrophilia (12). Of all cell classes, visualisation of eosinophils is the most affected by stain and preparation. Loss of defined granules can render them to look like a neutrophil due to the segmented nuclei. Both eosinophils and lymphocytes are also usually 1-5% of cells and disagreements in rare classes is punished hard by the F1 score. As lymphocytes are infrequent in sputum (19), error for this class, whether by human or SputNet, is likely to only minimally affect diagnoses of eosinophilic or neutrophilic inflammation.

Our study highlights the significant biases that are inherent to traditional human respiratory differential cell counts, specifically errors from counting only small subsets, subjective slide area selection, and natural human error. Our study also significantly differs from previous AI cell identification efforts by not only benchmarking against four independent readers, but also exploring the benefits of computational tools counting the whole slide. For the first time, we reconstructed the variability inherent that can occur in clinical cohort samples in standard 400-cell counts (9). The simulation shows how subset counts can misclassify eosinophilia and neutrophilia, particularly near diagnostic thresholds. A single 400-cell eosinophil count may therefore be least stable in the range where treatment decisions are made (11). Careful processing can reduce large compositional swings across a cytospin, and our respective laboratories in Perth and Newcastle have decades of experience in cytospins generation. Even with careful processing, the inherent randomness of cell deposition means that some fields of view will still sit below the slide mean. As SputNet or similar whole-slide methods can count the entire preparation in the time needed for a human subset count, they return a more complete estimate of cell proportions than a 400-cell subset count. Flow cytometry can also assess an entire respiratory sample, but it requires specialised infrastructure, panels (20), cost and expertise (21), which limits use in regional or primary care settings.

Induced sputum collection is well-established (22) and recommended by American Thoracic Society and European Respiratory Society guidelines. As mentioned in the introduction, integration of differential cell counts into routine care remains limited, largely because cytological analysis is time-intensive and expertise-dependent (3, 7, 8). New protein assays for eosinophilia and neutrophilia show promise (23-25) but are not yet validated for personalised clinical monitoring. Blood cell counts and FeNO often differ from airway cell counts, or fail to detect longitudinal changes in airway biology (26-28). Cytospin preparations remain amongst the lowest cost and rapid sample preparations, which makes a whole-slide cytological method easier to place in existing laboratories, research studies and trials. Combining this low barrier processing with a logged, whole-slide count by AI could support more consistent phenotyping in asthma, COPD and other chronic lung diseases.

Given that existing pathology centres are already prepared for sputum sample processing and are increasingly adopting digital microscopy (29), respiratory medicine should be seeking to leverage this infrastructure through technologies like SputNet. Patients are likely to be receptive to tools with significant potential benefit to chronic respiratory disease management, patient outcomes and clinical efficiency. A major use case is severe asthma and COPD, where routine sputum collection and treatable traits are already established (30, 31). In asthma alone, sputum immunophenotyping has been calculated to realise incremental cost-effectiveness of $3566 per quality-adjusted life year over FeNO (5, 32). Widespread adoption of AI-based technologies like SputNet would facilitate larger, more comprehensive cost-benefit analyses for sputum-guided disease management and strengthen statistical power in clinical trials.

This study has several limitations. Our dataset originated from three Australian research centres and we only utilised a single digital scanner for imaging. We also did not perform a priori sample size calculations. However, our novel data on annotator variability and cell class imbalance will inform future sample size calculations (33). Other deep learning detectors for BAL cytology have reported high accuracy (34-37). We did not compare SputNet against these other approaches as they were evaluated on different sample types, cell classes and metrics, making a direct comparison not possible. Although SputNet was evaluated on a locked test set that was never used for training or model selection, the test cytospins were drawn from the same centres and scanner as the development data. Broader dataset training and external testing will therefore be essential to identify weaknesses in performance that affect SputNet’s pathway to clinical translation. This will require testing against real-world heterogeneity across centres in stains, magnifications, debris profiles, cell densities, and scanners. Fortunately, cytospin preparations are robust for long-term archiving (38) and there undoubtedly exists a substantial global resource of historical respiratory studies for this purpose. These resources will also help to refine immunophenotype thresholds and improve prognostic utility in respiratory disease.

Another limitation is that we did not have access to clinical outcomes or treatment response data to validate SputNet against. SputNet was also not trained for identifying free eosinophil granules, a marker that is gathering clinical attention for its potential to identify active eosinophilic inflammation (31). This was outside the training of our annotators but is a further improvement that could be achieved through collaboration Beyond granules, the same approach could be trained on morphological features already proposed as clinically relevant, including variation in macrophage, neutrophil and eosinophil morphology (23, 39, 40) and macrophage content such as carbon or silica (41).

In summary, SputNet demonstrates how AI-based cytology tools can provide faster, more consistent, and potentially more accurate results than current manual counting approaches. Further validation and multicentre external validation of SputNet is needed, followed by implementation trials prior to any integration into clinical practice. Automated deep learning-based image analysis tools are already used across various other medical disciplines, including cardiology, ophthalmology, and gastroenterology. We propose that respiratory medicine also embraces this opportunity in respiratory sample cell counts.

## Supporting information

TRIPOD+AI

## Data Availability

All data produced in the present study are available upon reasonable request to the authors.

## Acknowledgements

We gratefully acknowledge Ms Sarada Lee, Mr Luke Berry, Mrs Kellie Fakes, and Mr Craig Schofield for data generation support, as well as the study participants and investigators who provided archived slides utilised in this research. A survey for physicians to provide their attitudes towards sputum counts and digital assessment methods can be found here: https://redcap.telethonkids.org.au/redcap/surveys/?s=P7AKNH3LJX4M7NW7.

## Funding

This project and L.W.G. were funded by the Western Australian (WA) Future Health Research and Innovation (FHRI) Fund (award number WANMA/IG2021/3). The funder had no role in study design, data collection, analysis, interpretation, or the decision to submit the manuscript

## Data Availability Statement

De-identified tile images and annotations, the analytical code and the trained models will be made available on reasonable request to the corresponding authors, subject to a data-sharing agreement.

## Conflict of Interest

The authors have no conflicts of interest to declare.

## Notes

### Competing Interest Statement

The authors have declared no competing interest.

### Author Declarations

Respiratory sample cytospins were sampled from archives of four research studies and collated imaging and analysis performed in this study under ethical approval from the University of Western Australia Human Research Ethics Committee, approval 2023/ET000729. Additional consent was not sought as these archived samples were all consented at the time of collection for the quantification of immune cells by differential count and participants consented to use in future research.

