## supplementary material for "SputNet: expert-level deep learning differential cell counts across whole respiratory cytospins expose the limits of manual counting"

ONLINE SUPPLEMENT

SputNet: whole-slide deep learning differential cell counts of respiratory cytospins and the sampling error of manual counting

Authors: Shayan Rasaratnam^1^, Gökhan Yildirim^2^, Yuliya V Karpievitch^3,4^, Jodie L Simpson^5^, Luke W Garratt^1,3,6^

^1^School of Medicine, University of Western Australia, Crawley, Western Australia

^2^Canva, Sydney, New South Wales

^3^Wal-yan Respiratory Research Centre, The Kids Research Institute of Australia, Crawley, Western Australia

^4^School of Biomedical Sciences, University of Western Australia, Crawley, Western Australia

^5^School of Biomedical Sciences and Pharmacy, University of Newcastle, Callaghan, New South Wales

^6^Institute for Respiratory Health, Nedlands, Western Australia

### Methods

#### Cytospin preparation

Respiratory sample cytospins were sampled from archives of four research studies across asthma, COPD and CF (1-4).

For sputum sample cytospins, these were prepared in a protocol similar to that published by Pizzichini et al (5). Patients were induced and sputum assessed for muco-cellular clumps, which were selected and dispersed using dithiothreitol (DTT) (3). Following DTT treatment, the sample was filtered, and the cell fraction was obtained by centrifugation. Specific details can be found in the original citations.

For bronchoalveolar lavage cytospins, samples solely underwent centrifugation to obtain the cellular fraction (6). For all samples, centrifuged cell pellets were resuspended, total cell counts were performed and cytospins prepared by centrifugation. Prepared cytospins were stained with either May-Grunwald Giemsa or Kwik-Diff™ and mounted with a coverslip.

#### Dataset splits

The development set consists of 3,678 different 1024x1024 100x magnification images of cytospins. For cross-validation the development set was divided into four folds grouped by cytospin, so that all images from one cytospin were assigned to the same fold; the folds contained 880, 980, 878 and 940 images.

The test set consists of 600 different 1024x1024 100x magnification images of cytospins. The counts of each annotator are as follows:

|  | Annotator A | Annotator B | Annotator C | Annotator D |
| --- | --- | --- | --- | --- |
| Eosinophil | 243 | 167 | 221 | 147 |
| Lymphocyte | 98 | 290 | 304 | 119 |
| Macrophage | 3,223 | 3,212 | 2,334 | 4,889 |
| Neutrophil | 3,996 | 4,106 | 3,324 | 4,086 |
| Columnar Epithelial | 197 | 728 | 242 | 65 |
| Squamous Epithelial | 234 | 332 | 208 | 67 |
| Unknown Cell | 11 | 23 | 411 | 151 |

#### Cell detection model

Cell detection was performed using Faster R-CNN (7) with a ResNet-50-FPN backbone, with a single modification to the model architecture by changing the number of outcome classes to two. One class to indicate a cell, and another class to indicate background (no cell). Starting from the pretrained weights for the COCO Dataset (8), the model was trained in two phases. In the head phase only the box predictor was trainable (learning rate 9.1 x 10^-4^) for up to six epochs; all layers were then unfrozen and fine-tuned (learning rate 1.0 x 10^-5^) for up to 20 epochs. In cross-validation, training stopped after four epochs without improvement in validation F1 and the best epoch was restored; the final model, trained on all development images without a validation fold, used the upper-median epoch counts from the four folds (three head epochs and 11 fine-tuning epochs).

Faster R-CNN outputs a list of bounding boxes, classes, and probabilities. A probability threshold is then used to filter bounding boxes, then boxes that overlap are also filtered to generate the bounding boxes that go to the classification model. Augmentations used for Faster R-CNN include random vertical flip, random horizontal flip, and random rotation. The Adam optimiser was used, and the batch size was set to one image. The loss function used was the standard multi-task loss for Faster R-CNN as implemented by Torchvision (9).

#### Classification model

Classification was generated using an EfficientNetV2-L architecture (10), starting from pretrained weights for the ImageNet challenge (11). The input of this model is a fixed 202 x 202 pixel field of view centred on each cell detected by the Faster R-CNN model; during training the crop is centred on the annotated cell centre with up to 16 pixels of random jitter. The output of this model is six possible cell classes: eosinophil, macrophage/monocyte, lymphocyte, neutrophil, columnar epithelial, squamous epithelial. The model was trained in two phases: the classification head alone (learning rate 9.1 x 10^-4^) for up to ten epochs, followed by fine-tuning of all layers (learning rate 1.0 x 10^-5^) for up to 30 epochs. In cross-validation, validation macro-F1 selected the best epoch and stopped training after four epochs without improvement; the final model, trained on all development cells without a validation fold, used the upper-median epoch counts from the four folds (six head epochs and 11 fine-tuning epochs). Augmentations used for EfficientNetV2-L included: trivial augment wide, random vertical flips, random horizontal flips, and crops to 60 pixel x 60 pixel, 104 pixel x 104 pixel, 122 pixel x 122 pixel, and 202 pixel x 202 pixel. Cross entropy loss weighted by normalised inverse class frequency, capped at 20 times the smallest class weight, was used with the Adam optimiser, and the batch size was set to 32.

For both Faster R-CNN and EfficientNetV2-L, hyperparameter sweep was performed across different learning rates, weight decay, and number of epochs to determine the best performance for the development set. The implementation was completed using the PyTorch library 2.13.0 and TorchVision 0.28.0 in Python 3.14.6 with CUDA, run as Slurm batch jobs on NVIDIA A100 GPUs.

All model development was performed by cross-validation within the development set. The 600-image test set was held out from every stage of development and was used once, with four independent cytologist annotation sets, to quantify inter-annotator agreement and to compare SputNet against each annotator.

#### Heterogeneity analyses

Count data generated from all cytospins in the study by SputNet was imported into RStudio™ (2026.01.0 Build 392) (12) and analysed using packages spatstat v. 3.5.1 (13), spdep v. 1.4.1 (14), stats v. 4.5.2 (15).
